# The T-cell clonal response to SARS-CoV-2 vaccination in inflammatory bowel disease patients is augmented by anti-TNF therapy and often deficient in antibody-responders

**DOI:** 10.1101/2021.12.08.21267444

**Authors:** Dalin Li, Alexander Xu, Emebet Mengesha, Rebecca Elyanow, Rachel M. Gittelman, Heidi Chapman, John C. Prostko, Edwin C. Frias, James L. Stewart, Valeriya Pozdnyakova, Philip Debbas, Angela Mujukian, Arash A Horizon, Noah Merin, Sandy Joung, Gregory J. Botwin, Kimia Sobhani, Jane C. Figueiredo, Susan Cheng, Ian M. Kaplan, Dermot P.B. McGovern, Akil Merchant, Gil Y. Melmed, Jonathan Braun

## Abstract

**Background:** Vaccination against SARS-CoV-2 is a highly effective strategy to protect against infection, which is predominantly mediated by vaccine-induced antibodies. Postvaccination antibodies are robustly produced by those with inflammatory bowel disease (IBD) even on immune-modifying therapies but are blunted by anti-TNF therapy. In contrast, T-cell response which primarily determines long-term efficacy against disease progression,, is less well understood. We aimed to assess the post-vaccination T-cell response and its relationship to antibody responses in patients with inflammatory bowel disease (IBD) on immune-modifying therapies.

**Methods:** We evaluated IBD patients who completed SARS-CoV-2 vaccination using samples collected at four time points (dose 1, dose 2, 2 weeks after dose 2, 8 weeks after dose 2). T-cell clonal analysis was performed by T-cell Receptor (TCR) immunosequencing. The breadth (number of unique sequences to a given protein) and depth (relative abundance of all the unique sequences to a given protein) of the T-cell clonal response were quantified using reference datasets and were compared to antibody responses.

**Results:** Overall, 303 subjects were included (55% female; 5% with prior COVID) (Table). 53% received BNT262b (Pfizer), 42% mRNA-1273 (Moderna) and 5% Ad26CoV2 (J&J). The Spike-specific clonal response peaked 2 weeks after completion of the vaccine regimen (3- and 5-fold for breadth and depth, respectively); no changes were seen for non-Spike clones, suggesting vaccine specificity. Reduced T-cell clonal depth was associated with chronologic age, male sex, and immunomodulator treatment. It was preserved by non-anti-TNF biologic therapies, and augmented clonal depth was associated with anti-TNF treatment. TCR depth and breadth were associated with vaccine type; after adjusting for age and gender, Ad26CoV2 (J&J) exhibited weaker metrics than mRNA-1273 (Moderna) (p=0.01 for each) or BNT262b (Pfizer) (p=0.056 for depth). Antibody and T-cell responses were only modestly correlated. While those with robust humoral responses also had robust TCR clonal expansion, a substantial fraction of patients with high antibody levels had only a minimal T-cell clonal response.

**Conclusion:** Age, sex and select immunotherapies are associated with the T-cell clonal response to SARS-CoV-2 vaccines, and T-cell responses are low in many patients despite high antibody levels. These factors, as well as differences seen by vaccine type may help guide reimmunization vaccine strategy in immune-impaired populations. Further study of the effects of anti-TNF therapy on vaccine responses are warranted.

## Introduction

Vaccination with mRNA or vector vaccines is immunogenic for SARS-CoV-2 and protective for occurrence and severity of COVID-19. Anti-SARS-CoV-2 antibodies dominate protection against initial infection ^1, 2^, whereas T-cells play a larger role in preventing disease progression ^3, 4^. The T-cell clonal response to SARS-CoV-2 vaccines in immunologically impaired individuals is poorly understood, as are effects of risk-factors on this aspect of the vaccine response. Here, a cohort of inflammatory bowel disease (IBD) patients are assessed for their clonal T-cell vaccine response, and its alteration by demographic factors and immunotherapy.

## Methods

The TCR clonal response to SARS-CoV-2 vaccines was assessed in 303 individuals with IBD, enrolled in a prospective registry at Cedars-Sinai between January and June 2021. Samples were collected longitudinally at the time of dose 1, dose 2, and 2 and 8 weeks after dose 2.

### Subjects

Inflammatory bowel disease patients (N=303) were recruited in Los Angeles, CA, USA between January and June 2021 under the CORALE-IBD protocol approved by the Cedars Sinai Institutional Regulatory Board. Details of this cohort were recently reported ^5, 6^. Participants completed baseline surveys detailing demographics and medical history at the time of vaccination, and were offered blood sampling after dose 1 (from 5 days after dose 1 until the day of dose 2), after dose 2 (from 2 to 13 days after dose 2), and at 2 weeks (14 to 29 days after dose 2), and 8 weeks (30 to 84 days after dose 2). Prior COVID-19 status was defined by positive IgG(N) at any timepoint, or individuals with a prior clinical diagnosis of COVID-19. COVID-19 experienced individuals were excluded from analysis except where specifically noted. Most participants received mRNA vaccines, and except where indicated, analysis was restricted to this subgroup.

### Antibody assessment

Plasma antibodies to the receptor binding domain of the S1 subunit of the viral spike protein [IgG(S-RBD)] were quantified using the SARS-CoV-2 IgG-II assay (Abbott Labs, Abbott Park, IL). as previously described ^5^.

### T cell clonal analysis

Immunosequencing of the CDR3 regions of human TCRβ chains was performed on blood genomic DNA using the immunoSEQ Assay (Adaptive Biotechnologies), which includes bias-controlled multiplex PCR, high-throughput sequencing, and identification and quantitation of absolute abundance of unique TCRβ CDR3 regions, and quantitation of the corresponding T cell fractions by template count normalization^7^. Attribution of TCR sequences to SARS-CoV-2 spike or other non-spike SARS-CoV-2 protein specificities were assigned as described by Alter et al. and Sinder et al. ^8, 9^. The breadth summary metric was calculated as the number of unique annotated rearrangements among total number of unique productive rearrangements in the individual sample’s dataset. The depth metric was calculated by combining two elements; (a) the raw frequency of each rearrangement in the total repertoire in the individual sample’s dataset, and an estimate of clonal generations of the lineage represented by each rearrangement. The resultant depth metric estimates the relative number of clonal expansion generations across the TCRs, normalized by the total number of TCRs sequenced in the sample. Hence, the metric can range from negative to positive values^9^.

### Data analysis

Comparison of TCR breadth and depth used Mixed Linear Model across time points and Generalized Linear Model within time points. Where possible, inverse normal transformation was performed, and age and sex were included as covariates. Confidence intervals for binomial probabilities were computed using exact methods. Geometric means and confidence intervals were calculated for the log-transformed antibody levels. Other analyses are specified in the individual figures. Analyses were restricted to individuals with mRNA vaccines and no prior COVID-19 experience unless stated otherwise.

### Data availability

Requests for de-identified data may be directed to the corresponding authors (J.B., G.M.) and will be reviewed by the Office of Research Administration at Cedars-Sinai Medical Center before issuance of data sharing agreements. Data limitations are designed to ensure patient and participant confidentiality.

## Results

Demographics and clinical metadata are summarized in Table 1. The T-cell clonal response to vaccination across different time points is shown in Figure 1A. At dose 1, spike-specific breadth and depth of SARS-CoV-2 clones were low (reflecting their basal level in an individual’s repertoire). Levels peaked two weeks post second vaccination (P=4.64E-25 and 2.42E-25 relative to dose 1 levels, for breadth and depth, respectively). From this peak, levels declined at 8 weeks post second vaccination but were still significantly elevated (relative to dose 1, 1.08E-11 and 5.30E-14, for breadth and depth respectively). In contrast, no changes were observed in T-cell clonal metrics for non-spike clones, demonstrating the specificity of the vaccine responses.

|  | Total | Dose 1 | Dose 2 | 2 Weeks | 8 Weeks |
| --- | --- | --- | --- | --- | --- |
| n | 303 | 110 | 158 | 153 | 184 |
| race, n(%) |  |  |  |  |  |
| Asian | 7(2.36) | 2(1.92) | 3(1.94) | 6(3.97) | 3(1.64) |
| Black or African American | 5(1.68) | 2(1.92) | 3(1.94) | 4(2.65) | 2(1.09) |
| Multiple | 4(1.35) | 2(1.92) | 2(1.29) | 2(1.32) | 3(1.64) |
| Other | 10(3.37) | 3(2.88) | 2(1.29) | 6(3.97) | 8(4.37) |
| Prefer not to answer | 3(1.01) | 1(0.96) | 2(1.29) | 2(1.32) | 1(0.55) |
| White | 268(90.24) | 94(90.38) | 143(92.26) | 131(86.75) | 166(90.71) |
| Hispanic , n(%) | 15(5.05) | 7(6.73) | 8(5.16) | 9(5.96) | 9(4.92) |
| Gender, female n(%) | 166(55.89) | 58(55.77) | 88(56.77) | 80(52.98) | 106(57.92) |
| Vaccine type, n(%) |  |  |  |  |  |
| BNT162 (Pfizer/BioNtech) | 160(52.81) | 67(60.91) | 90(56.96) | 79(51.63) | 97(52.72) |
| JNJ-78436725 (Johnson & Johnson) | 15(4.95) | 9(8.18) | - | 9(5.88) | 13(7.07) |
| mRNA-1273 (Moderna/NIH) | 128(42.24) | 34(30.91) | 68(43.04) | 65(42.48) | 74(40.22) |
| Prior COVID-19 History, n(%) | 15(5.08) | 6(5.88) | 6(3.9) | 5(3.33) | 6(3.3) |
| Treatments, n(%) |  |  |  |  |  |
| No Immune suppression | 48(16.22) | 15(14.02) | 28(18.18) | 22(14.57) | 29(16.11) |
| Anti-TNF | 104(35.14) | 35(32.71) | 54(35.06) | 54(35.76) | 65(36.11) |
| Other biologics (anit-IL23, anti-integrin) | 126(42.57) | 48(44.86) | 64(41.56) | 66(43.71) | 75(41.67) |
| Immunomodulators | 18(6.08) | 9(8.41) | 8(5.19) | 9(5.96) | 11(6.11) |
| COVID-19 TCR matrices, mean(s.d.) |  |  |  |  |  |
| clonal breadth | 2.05e-04(1.42e-04) | 1.24e-04(1.26e-04) | 2.03e-04(1.55e-04) | 2.87e-04(1.51e-04) | 1.93e-04(9.64e-05) |
| clonal depth | 64.26(84.19) | 22.26(49.83) | 76.13(111.82) | 102.45(92.14) | 50.71(47.47) |
| clonal breadth, Spike only | 4.49e-05(5.53e-05) | 2.29e-05(2.99e-05) | 5.04e-05(6.74e-05) | 7.69e-05(6.43e-05) | 4.35e-05(3.73e-05) |
| clonal depth, spike only | 2.06(29.04) | -10.59(12.89) | 5.86(41.77) | 13.91(30.94) | -2.66(15.18) |
| Age group, n(%) |  |  |  |  |  |
| <=30 | 44(14.52) | 16(14.55) | 28(17.72) | 23(15.03) | 23(12.5) |
| 30-40 | 83(27.39) | 31(28.18) | 41(25.95) | 48(31.37) | 44(23.91) |
| 40-50 | 71(23.43) | 30(27.27) | 38(24.05) | 38(24.84) | 36(19.57) |
| 50-60 | 45(14.85) | 14(12.73) | 24(15.19) | 25(16.34) | 30(16.3) |
| >60 | 60(19.8) | 19(17.27) | 27(17.09) | 19(12.42) | 51(27.72) |

**Figure 1.**
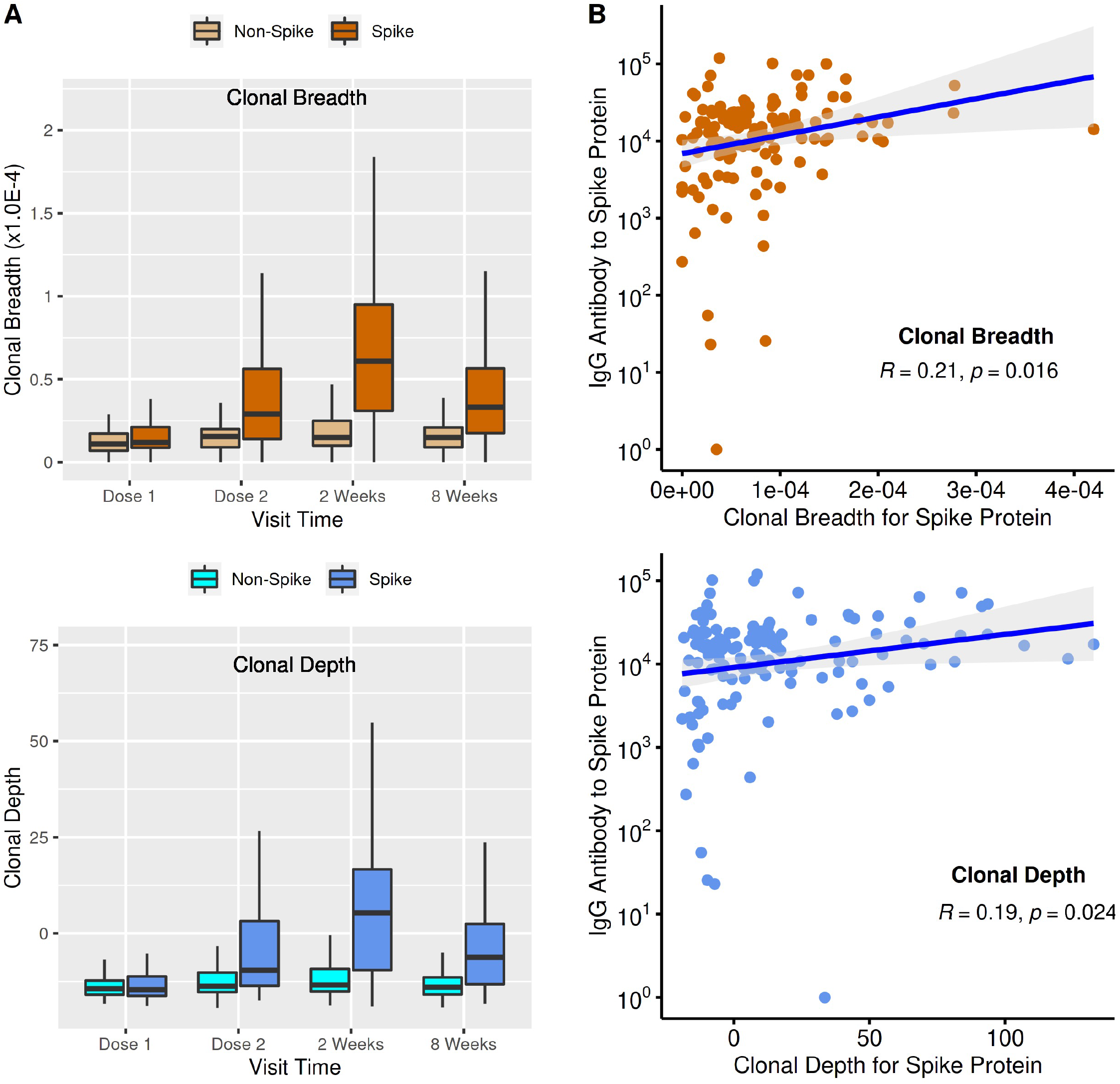
T-cell clonal response and antibody levels to SARS-CoV-2 immunization. (A) T-cell clonal response to SARS-CoV-2 vaccination. Box plots show mean, quartiles, and data range. Relative to dose 1, p values (mixed-effect model analysis with adjustment for age and sex) for dose 2, 2 weeks post 2^nd^ vaccination, and 8 weeks post 2^nd^ vaccination were: breadth (1.04E-8, 4.64E-25,1.08E-11); depth (9.87E-11, 2.42E-25,5.30E-14). (B) Comparison of T-cell clonal response metrics to anti-spike IgG levels (Spearman’s Correlation).

Spike-specific T-cell and antibody responses were compared at week 2 post dose 2, which corresponds to the peak of both antibody and T-cell vaccine responses ^10-12^ (Figure 1B). The two responses were significantly but only moderately correlated (R = 0.19 to 0.21). Among those with low antibody response, T-cell clonal breadth and depth were low, suggesting that those with impaired humoral vaccine response have similarly impaired cellular responses. However, among individuals with the lowest T-cell response, the majority discordantly had moderate or high antibody levels.

The spike-specific clonal breadth was preserved across age groups, but clonal depth reduced substantially with age (Figure 2A, P=3.62E-4 for trend test). There was no statistically significant association between sex and spike T-cell clonal responses at 2 weeks after dose 2 (eFigure 1). However, at 8 weeks the T-cell clonal response was increased in females versus males (P=0.083 and 0.0077, for breadth and depth respectively).

**Figure 2.**
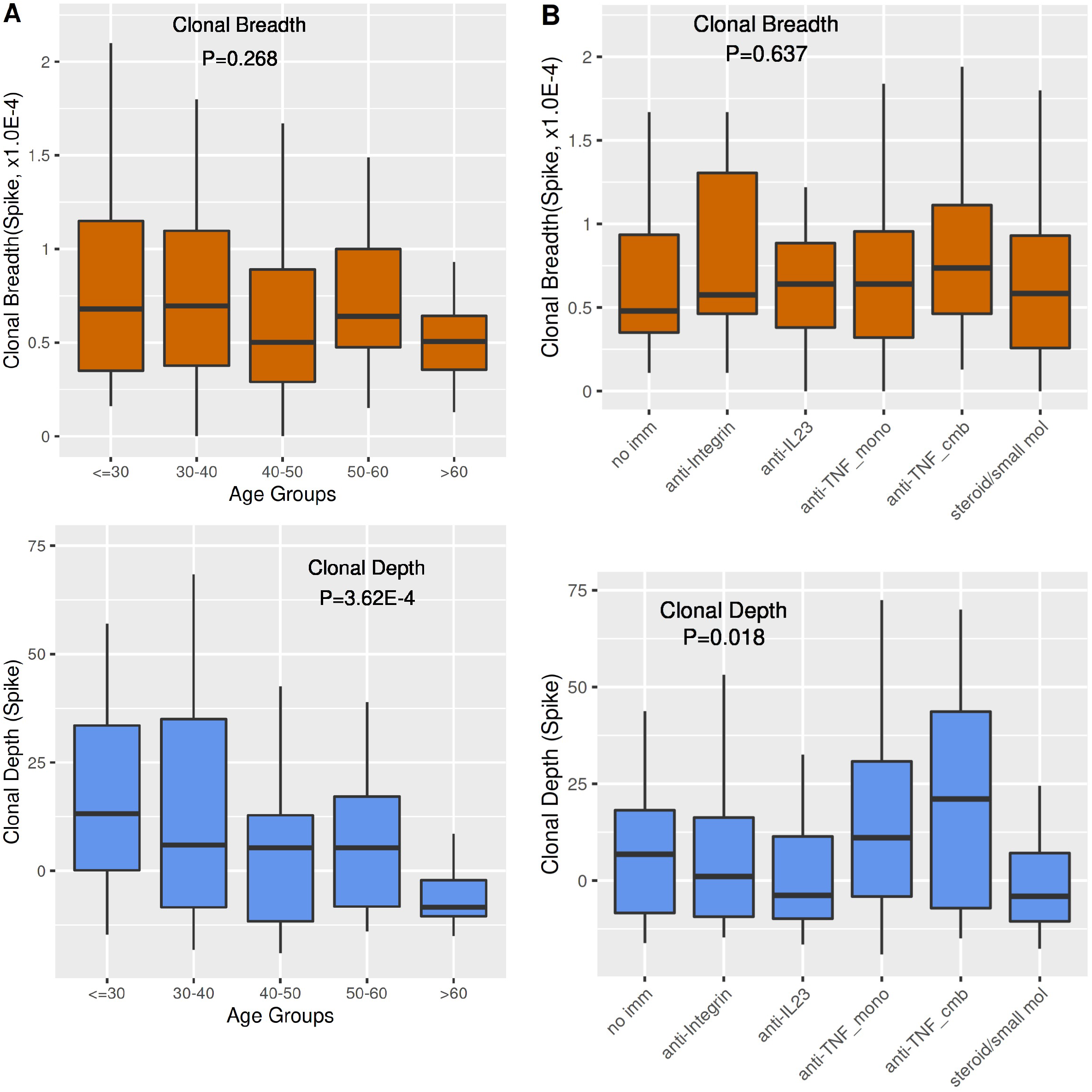
Effect of age and immunologic treatment on T-cell clonal response. (A) Age. Numbers of subjects by age group are tabulated in Table 1. (B) Immunologic treatment. No Imm (no treatment, 5-aminosalicylates, rectal steroids; N=19), anti-Integrin (N=14), anti-IL23 (N=36), anti-TNF_mono (monotherapy with anti-TNF, N=36), anti-TNF_cmb (Combined therapy with anti-TNF and a thiopurine or methotrexate, N=11), steroids/small mol (systemic corticosteroids, or monotherapy with thiopurines, methotrexate, or Janus kinase (JAK) inhibitors, N=16). Boxes are mean value, bars are data range, and p-values were calculated by ANOVA after adjustment for age, sex, vaccine type and COVID history.

IBD disease type (Crohn’s disease vs. ulcerative colitis) had minimal effects on the temporal kinetics or levels of spike T-cell clonal response to vaccines (eFigure 2). T-cell clonal depth was significantly but selectively affected by suppressive immunotherapy (Figure 2B, ANOVA p= 0.018). There were no significant effects of anti-IL12/23, anti-integrin, or steroids/small molecular treatments in comparison to patients with no immune treatments. Interestingly, we observed an augmentation with anti-TNF (p= 0.0174) after adjustment for age and sex, with consistent trends in anti-TNF monotherapy or in combination with immunomodulators.

No significant differences were observed between the T-cell clonal responses to the two mRNA vaccines assessed in this cohort at 2 weeks after dose 2, although a marginal difference was observed at 8 weeks for clonal breadth favoring mRNA-1273 (P=0.047, eFigure 3). Compared to mRNA vaccination, Ad26.COV2.S induced a smaller spike T-cell clonal response at both 2 weeks and 8 weeks after the single vaccination dose.

As expected, COVID-19 experienced subjects at dose 1 had significantly increased clonal T-cell breadth and depth compared to COVID-19 naïve subjects (eFigure 4). However, no significant differences were observed between experienced and naïve subjects in the peak TCR response (2 weeks).

## Discussion

This study assesses the T-cell clonal response to SARS-CoV-2 vaccine, to directly enumerate SARS-CoV-2 spike-specific T-cell clonal diversity (breadth) and clone size (depth) in immune-impaired individuals. Interrogation of our IBD patient cohort permitted assessment under select and discrete modes of therapeutic immunosuppression. Few studies have assessed the T-cell response to SARS-CoV-2 vaccines, and with few exceptions ^12^ have used methods that enumerate SARS-CoV-2-specific T-cells based on peptide-stimulated cytokine production ^10, 11, 13, 14^. Such studies don’t permit assessment of repertoire diversity and clonal size, important factors in protective T-cell immunity ^3, 4^.

Consistent with reported kinetics of polyclonal functional T-cell response to vaccination ^10-12^, T-cell clonal response peaked two weeks after the second vaccination dose. Although antibody response also peaks at 2 weeks ^5^, antibody levels provided limited predictiveness for the T-cell clonal response induced by vaccination, particularly for individuals with a low T-cell response. This is consistent with findings reported from polyfunctional T-cell assessment ^10, 11, 13, 14^. In the context of reimmunization strategies, T-cell assessment may be important to evaluate both initial vaccine response and persistence of immunity after vaccination ^15, 16^.

We observed that as age increased, clonal depth in T-cell response to COVID-19 vaccine decreased while clonal breadth was unaffected. This suggests that the potential spike-specific T-cell repertoire is maintained with age, but the burst size of the clonal response is curtailed, an observation previously reported in the global and influenza T-cell repertoire ^17, 18^. The T-cell clonal response was reduced 8 weeks post vaccination in males, mostly via the impact on clonal depth.

Immune-modifying therapy also reduced the T-cell response, again via its selective effect on clonal depth, and thus the capacity of potential clones to expand after vaccination. In contrast, the T-cell response was preserved with biologic therapies targeting IL12/23 and integrins, and paradoxically augmented by anti-TNF therapy. If confirmed, this may reflect a differential effect of anti-TNF therapy on T-cell clonal expansion and effector states besides cytokine production.

Taken together, these observations on age, sex, and immunotherapies have potential significance when considering groups to prioritize for SARS-CoV-2 reimmunization. We also observed suggestive signals for vaccine type on the T-cell clonal response, analogous to the reduced levels of antibody response with Ad26.SARS.CoV.2 in this same cohort ^5^. Due to the small number of Ad26.SARS.CoV.2 recipients studied, those differences should be interpreted with caution.

Limitations of this study include a cohort of only individuals with IBD, lack of racial diversity, and a tertiary center population, which reduce generalizability. Furthermore, direct TCR sequencing detects only a minor subset of index antigen-reactive clones among the much larger number of private clones ^7^.

## Conclusion

Age, sex and select immunotherapies might be associated with the T-cell clonal response to SARS-CoV-2 vaccines. A low T-cell response is poorly predicted by antibody levels.

## Data Availability

All data produced in the present study are available upon reasonable request to the authors.

## Acknowledgements

This study was supported by the Leona M. and Harry B. Helmsley Charitable Trust, the Widjaja Foundation Inflammatory Bowel and Immunobiology Research Institute, and the National Institute of Diabetes and Digestive and Kidney Disease Grants P01DK046763 and U01DK062413, the Cedars-Sinai Precision Health Initiative, the Erika J. Glazer Family Foundation, and through the Serological Sciences Network, grant NCI U54-CA260591.

We thank all those who have contributed to the CORALE-IBD Vaccine study: James Beekley, Sarah Contreas, Joseph Ebinger, Ergueen Herrera, Amy Hoang, Nathalie Nguyen, Sarah Sternbach, Nancy Sun, Min Wu, Keren Appel, Andrea Banty, Edward Feldman, Christina Ha, Dmitry Karayev, Benjamin Kretzman, Rashmi Kumar, Susie Lee, Shervin Rabizadeh, Theodore Stein, Gaurav Syal, Stephan Targan, Eric Vasiliauskas, David Ziring, Brigid Boland, Mary Hanna, Elizabeth Khanishian, Melissa Hampton, Justina Ibrahim, Ashley Porter, Shane White, Cindy Zamudio.

## Author Contributions

These authors equally contributed to the study: Dalin Li and Alexander Xu; Jonathan Braun, Dermot McGovern, Gil Melmed.

Acquisition of data: GM, JB, EM, DL, AX, GB, KS, AAH, HM, RE, RMG, HC, IMK, JCP, ECF, JLS

Analysis and interpretation of data: all authors

Drafting of the manuscript: DL, JB

Critical revision of the manuscript for important intellectual content: all authors

Statistical analysis: DL, AX

Obtained funding: GM, JB, DM, SC, JCF

Study supervision: GM, JB, DM, AM

---

**eFigure 1.**
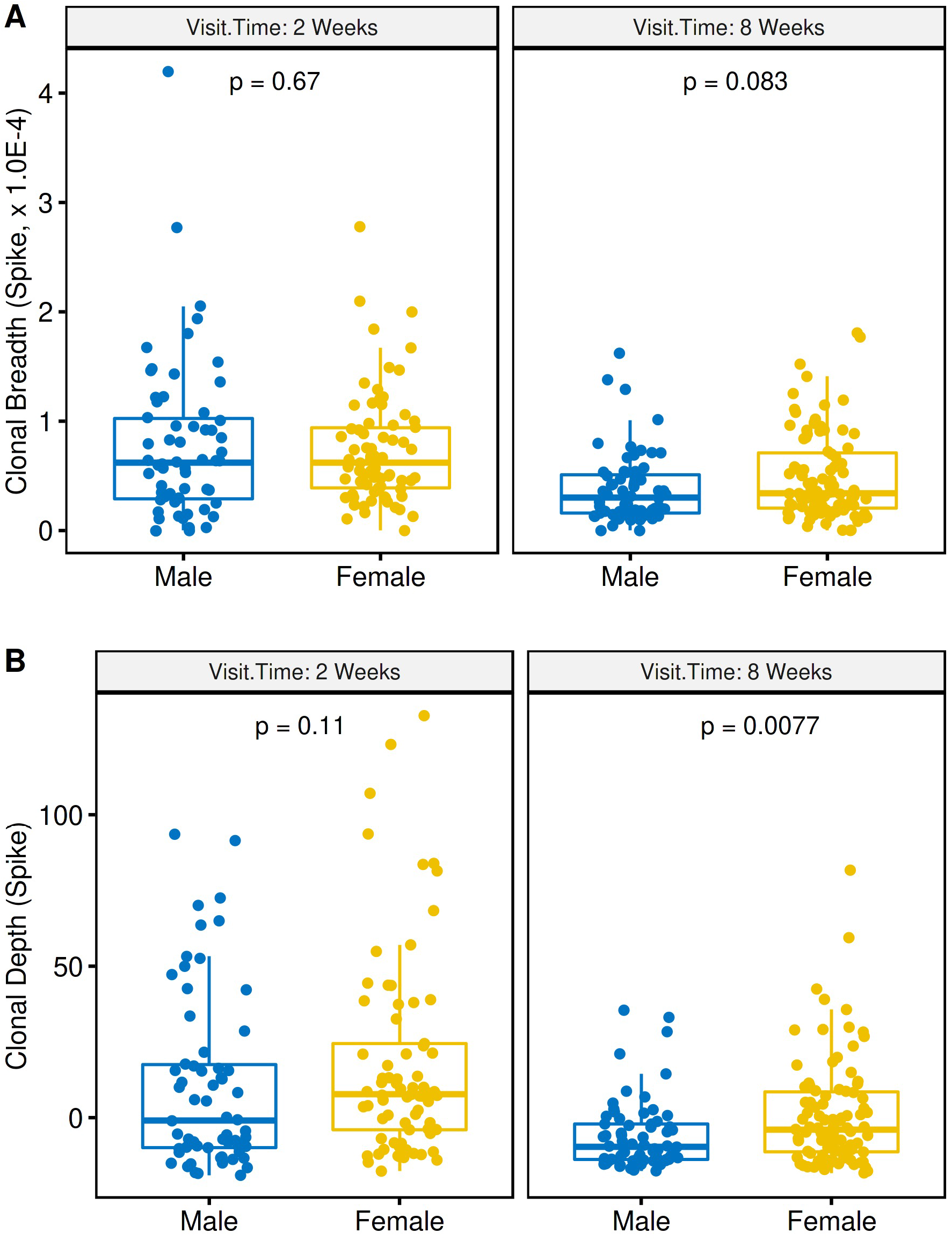
Effect of gender on spike T cell clonal response to vaccination. (A) T cell clonal breadth at week 2 (left) or week 8 (right) post second vaccination. (B) T cell clonal depth at week 2 (left) or week 8 (right) post second vaccination. Box plots show mean, quartiles, and data range. P values were calculated using a mixed-effects model (with adjustment for age and gender) comparing dose 1 to either 2 weeks post 2^nd^ vaccination (left) or 8 weeks (right) post 2^nd^ vaccination.

**eFigure 2.**
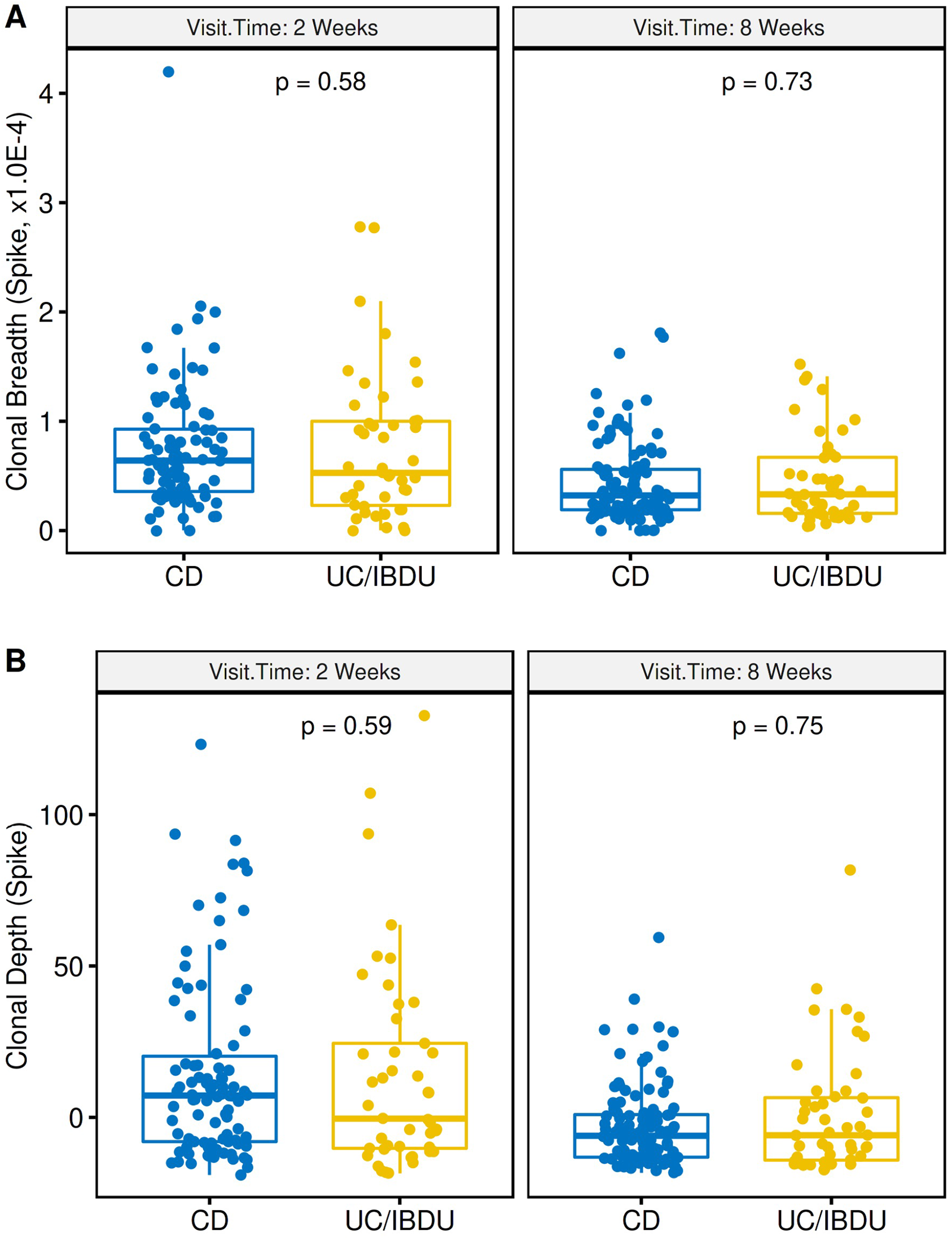
Effect of IBD disease type on spike T cell clonal response to vaccination. (A) T cell clonal breadth at week 2 (left) or week 8 (right) post second vaccination. (B) T cell clonal depth at week 2 (left) or week 8 (right) post second vaccination. CD, Crohn’s disease; UC/IC, Ulcerative colitis and indeterminant colitis. Box plots show mean, quartiles, and data range. P values are a mixed model analysis (with adjustment for age and gender) comparing dose 1 to either 2 weeks post 2^nd^ vaccination (left) or 8 weeks (right) post 2^nd^ vaccination.

**eFigure 3.**
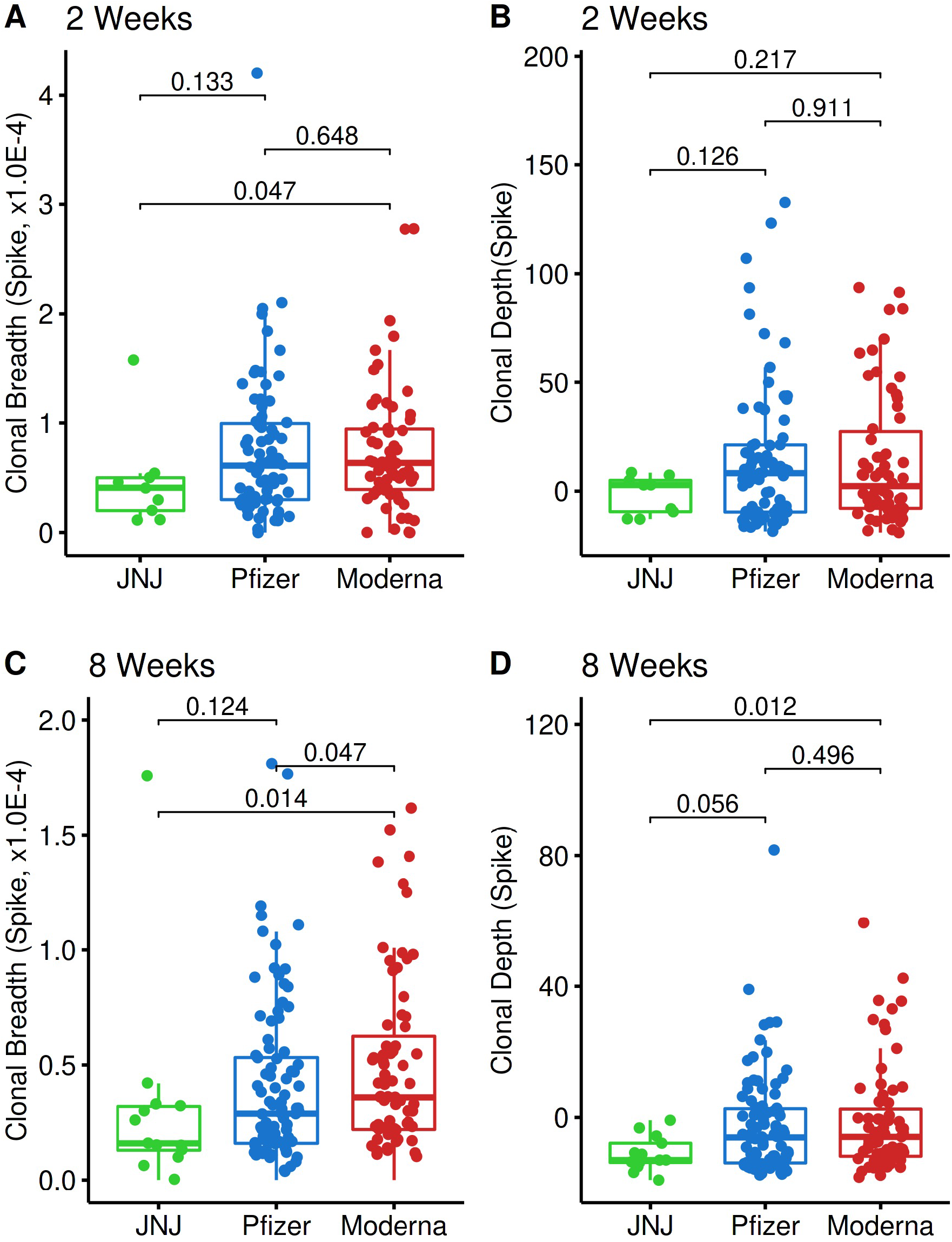
Vaccine type and the T cell clonal spike response. T cell clonal spike responses were tabulated 2 weeks (A and C) or 8 weeks (B and D) after completion of vaccination regimen (two doses for mRNA vaccines, one dose for the vector vaccine). Subject numbers were Pfizer (BNT162b2, N=160), Moderna (mRNA-1273, N=128), JNJ (Ad26.COV2.S, N=15). P values were calculated using a mixed-effects model (with adjustment for age and gender) for the indicated comparisons.

**eFigure 4.**
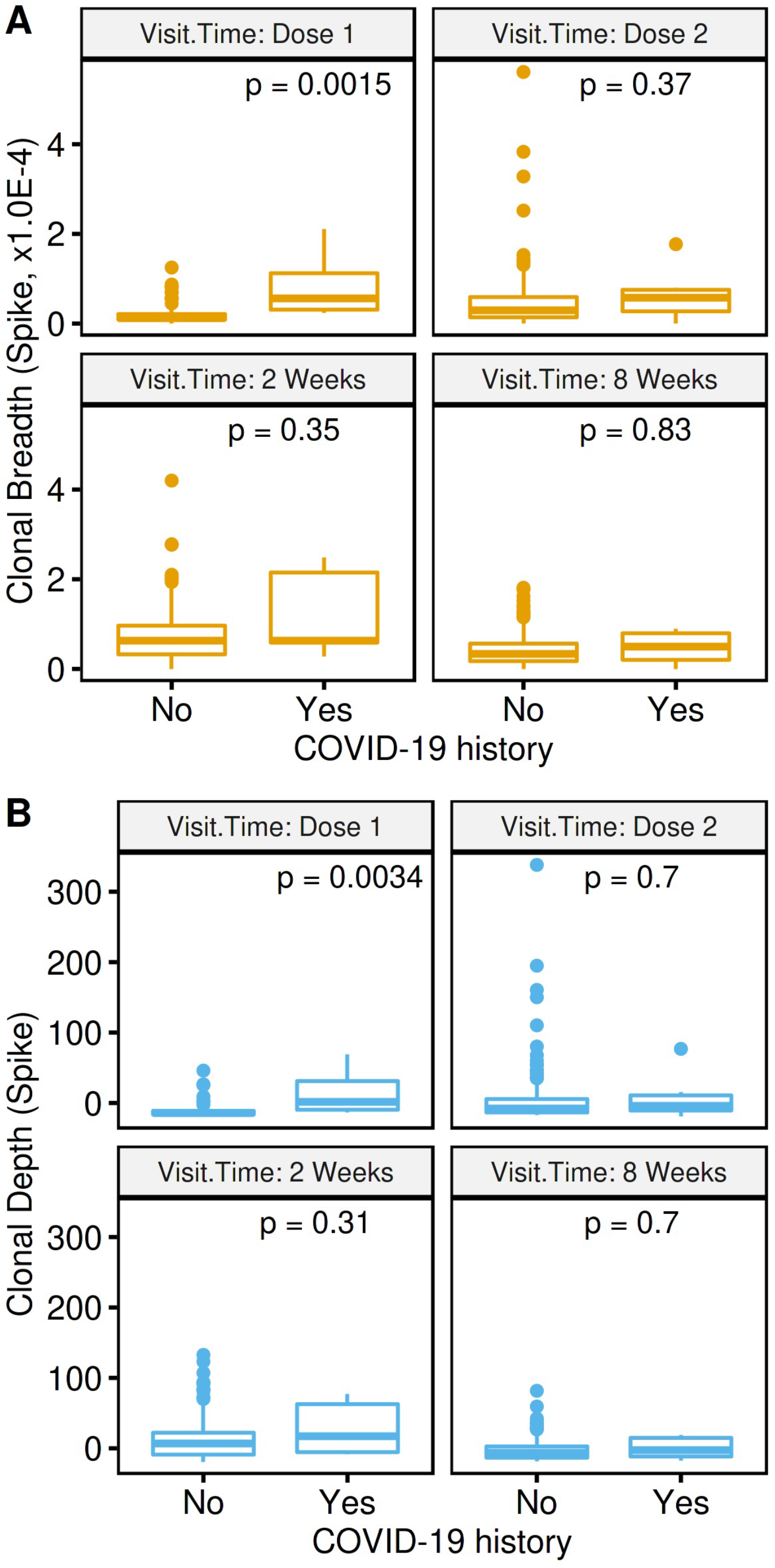
Effect of COVID-19 naïve and experienced status on T cell clonal response. The numbers of subjects were 288 (naïve) and 15 (experienced). (A) Dose 1; (B) Dose 2; (C) 2 weeks post second vaccination; (D) 8 weeks post second vaccination. Box plots show mean, quartiles, and data range. P values were generated from a mixed-effects model (with adjustment for age and gender) comparing naïve and experienced subjects.

